# Optimising the balance of acute and intermediate care capacity for the complex discharge pathway: computer modelling study during COVID-19 recovery in England

**DOI:** 10.1101/2021.10.25.21265475

**Authors:** Z Onen-Dumlu, AL Harper, PG Forte, AL Powell, M Pitt, C Vasilakis, RM Wood

## Abstract

**Objectives:** While there has been significant research on the pressures facing acute hospitals during the COVID-19 pandemic, there has been less interest in downstream community services which have also been challenged in meeting demand. This study aimed to estimate the theoretical cost-optimal capacity requirement for ‘step down’ intermediate care services within a major healthcare system in England, at a time when considerable uncertainty remained regarding vaccination uptake and the easing of societal restrictions.

**Methods:** Demand for intermediate care was projected using an epidemiological model (for COVID-19 demand) and regressing upon public mobility (for non-COVID-19 demand). These were inputted to a computer simulation model of patient flow from acute discharge readiness to bedded and home-based Discharge to Assess (D2A) intermediate care services. Cost-optimal capacity was defined as that which yielded the lowest total cost of intermediate care provision and corresponding acute discharge delays.

**Results:** Increased intermediate care capacity is likely to bring about lower system-level costs, with the additional D2A investment more than offset by substantial reductions in costly acute discharge delays (leading also to improved patient outcome and experience). Results suggest that completely eliminating acute ‘bed blocking’ is unlikely economical (requiring large amounts of downstream capacity), and that health systems should instead target an appropriate tolerance based upon the specific characteristics of the pathway.

**Conclusions:** Computer modelling can be a valuable asset for determining optimal capacity allocation along the complex care pathway. With results supporting a Business Case for increased downstream capacity, this study demonstrates how modelling can be applied in practice and provides a blueprint for use alongside the freely-available model code.

## Introduction

Many health and care systems have experienced unprecedented pressures as a result of the global COVID-19 pandemic [1]. With rapid transmission and accompanying societal restrictions, acute hospitals have had to contend with uncertain fluctuations in the nature and magnitude of demand for their services [2]. In response, there has been significant academic interest in forecasting the number of COVID-19 cases requiring hospitalisation; both to determine the extent of any mitigating societal restrictions (to prevent health and care systems from being overwhelmed), and to support effective local management of hospital capacity [3,4]. Yet, while secondary care has been directly exposed to the immediate effects of COVID-19, a proportion of admitted patients will require some form of ‘step-down’ intermediate care following their hospital discharge, and management of this capacity has received significantly less attention.

Effectively managing intermediate care capacity is especially important in times of high COVID-19 incidence. First, insufficient capacity puts severe pressure on upstream services, propagating ‘blockages’ in patient flow that, in turn, lead to increased stress on the (more expensive) acute care bed base, emergency department overcrowding, cancellation of elective procedures, and ambulance offload delays [5,6]. Second, delays to acute discharge are known to be associated with negative patient outcomes, including reduced physical and mental wellbeing and increased risk of nosocomial infections and cognitive decline [7]. Third, it is the same group of predominantly older individuals more predisposed to severe COVID-19 (and hospitalisation, especially prior to full vaccination) that are most likely to require intermediate care upon discharge [8]. Thus, with such high demand, the inability to ensure smooth patient flow from acute care carries the very real risk of destabilising the whole health and care system and evidence of this has already emerged during the pandemic [9,10].

The objective of this study was to estimate appropriate capacity requirement for intermediate care services within a major English healthcare system at a time of considerable uncertainty in the determinants of patient demand during the COVID-19 recovery period, namely vaccination uptake and the easing of societal restrictions. In assessing the most appropriate capacity requirement, consideration was given to the cost of both intermediate care service provision and the acute capacity required to support any delays in discharging patients (recognising that, in general, increases in the former would bring about reductions in the latter, and vice versa). The amount of intermediate care capacity yielding the lowest total cost is defined as the theoretical cost-optimal capacity requirement, whose estimate we sought by means of an empirically informed computer simulation study.

### Organisation of intermediate care services

In common with many health and care services around the world – particularly those following a European or North American model – intermediate care is used in the UK to ‘bridge’ the gap between acute hospital and the usual place of residence for individuals typically of older age and greater complexity [11]. The underlying rationale is to reduce unnecessary use of acute hospital resources beyond the point a patient is deemed medically fit for discharge by transferring them to a setting in which their longer-term health and social care needs can be properly assessed. In the English National Health Service (NHS), the Discharge to Assess (D2A) model [12], one of the relevant care system interventions introduced to tackle this problem, accounts for three time-limited pathways which provide ‘step-down’ care for a period of up to six weeks (Figure 1). Patients discharged on Pathway 1 (P1) return to their usual place of residence and receive domiciliary visits from community health services. If more intensive post-acute rehabilitation is required then patients – who are expected to return to their usual place of residence eventually – may be discharged on Pathway 2 (P2), which involves transfer to a non-acute bedded facility for up to six weeks. Pathway 3 (P3) is also non-acute bed-based care, but is reserved for those requiring the most complex health and social care need assessments. Many of the patients in this pathway will subsequently go on to a long-term care home placement. In England, of those that enter a D2A pathway following an acute admission, it is expected that at least 90% will require P1, with a maximum of 8% and 2% requiring P2 and P3 respectively [12].

**Figure 1.**
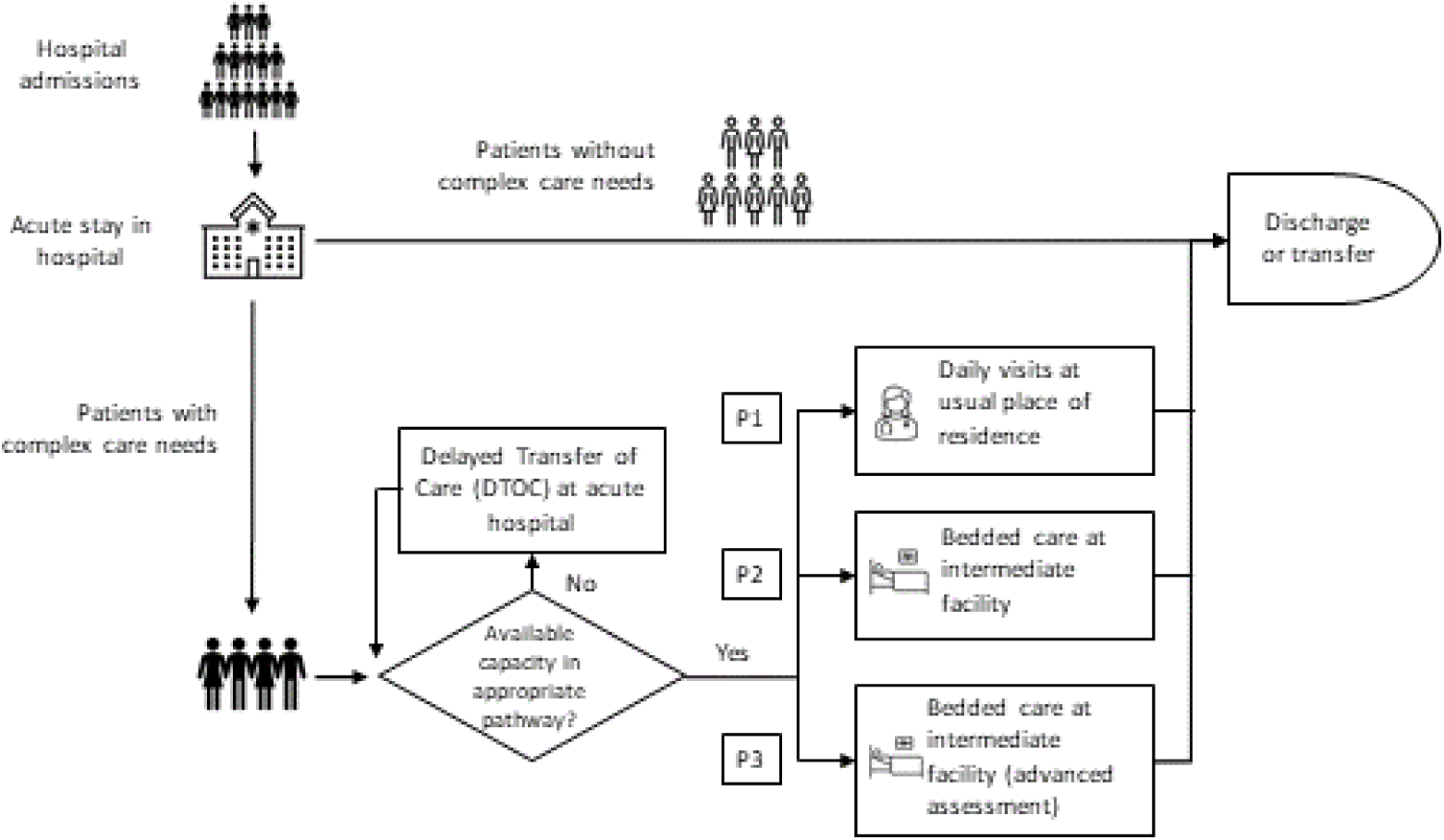
Organisation of intermediate care services in the English National Health Service (NHS).

### Review of computer modelling literature

Computer modelling has proved a useful resource in supporting effective healthcare management, with recent decades demonstrating a variety of influential case studies [13,14]. Much of the literature has focussed on improving the operation of individual acute services, such as forecasting emergency demand, scheduling outpatient consultations and surgery, and optimising intensive care capacity [15,16]. With regard to modelling community care, investigators have considered bedded [17] as well as home visits [18]. However, the authors of a review of modelling studies of community services [19] remarked that multiple care settings are rarely considered (and additionally that time-varying demand is not captured – an important property for modelling the effect of COVID-19 induced demand).

In the few modelling studies that do consider the interaction between acute and community care, the scope is often restricted to a specific type of disease, such as stroke care, childhood asthma, or other chronic diseases [20-21]. Yet these accounts do demonstrate the value in modelling the wider scope and context of the patient pathway, as opposed to specific parts of it. Without the ability to consider these interactions, it is more difficult to reliably assess the full extent of various capacity considerations in the performance and operation of upstream services, and derive a more holistic perspective of health and care system function.

## Methods

### Setting

The Bristol, North Somerset and South Gloucestershire (BNSSG) healthcare system covers a population of approximately one million individuals in South West England. Acute services are provided by two hospital trusts and intermediate care is provided through D2A pathways by a single community services provider. At the time of the study (May 2021), the D2A pathways had high occupancy rates with all 546 of the available daily home visits being used in the P1 pathway, 85% of the 177 P2 beds in use, and all 172 of the available P3 beds in use. This contributed to significant delays to accessing the three pathways, with 64, 62 and 83 patients in acute care ready and awaiting discharge (henceforth, this is referred to as the number of ‘blocked’ acute beds). Mean D2A lengths of stay were 13, 29 and 43 days for the P1-3 pathways, with respective standard deviations of 16, 22 and 31 days illustrating significant variation.

Around the time of the study, new COVID-19 hospitalisations were reducing from their peak on 21 January 2021 which had followed the imposition of a national lockdown on 5 January 2021. While acute COVID-19 bed occupancy in BNSSG was just 1.4% of its 439 peak, much uncertainty remained regarding the easing of societal restrictions (which began 8 March 2021 and were due to end by 21 June 2021), and the uptake of the vaccine (which started being administered in December 2020 and was due to be offered to all adults by 31 July 2021). Uncertainty also existed around the ability to reduce D2A lengths of stay to their target levels from actual durations that were considered to be too high and unaffordable in the longer term. For the BNSSG healthcare system, the concern was how these three aspects of uncertainty would affect future intermediate care demand, and thus determine appropriate capacity requirement.

### Scenario analysis

Scenario analysis was performed in order to examine the sensitivity of modelled capacity requirement to the above-mentioned aspects of uncertainty (Table 1).

**Table 1.** Scenarios considered in this study, with baseline values are indicated by (*). Note that while the mean is reported here, the full distribution of length of stay has been used in the model (to capture the aforementioned significant variation).

| Scenario | Vaccination uptake | End to societal restrictions | Mean length of stay in days (P1, P2, P3) |
| --- | --- | --- | --- |
| 1 | 90% * | 21 June 2021 * | 13, 29, 43 * |
| 2 | 90% * | 21 June 2021 * | 10, 21, 28 |
| 3 | 90% * | 21 August 2021 | 13, 29, 43 * |
| 4 | 90% * | 21 August 2021 | 10, 21, 28 |
| 5 | 75% | 21 June 2021 * | 13, 29, 43 * |
| 6 | 75% | 21 June 2021 * | 10, 21, 28 |
| 7 | 75% | 21 August 2021 | 13, 29, 43 * |
| 8 | 75% | 21 August 2021 | 10, 21, 28 |

On 22 February 2021, the UK government published its ‘roadmap’ for the easing of societal restrictions, with four incremental relaxations to take effect no sooner than 8 March, 12 April, 17 May and 21 June [22]. At the time of the study, the first three stages had proceeded as planned, although concerns around transmission of the ‘Delta’ SARS-CoV-2 variant had raised the prospect of postponing the final removal of restrictions [23]. Accordingly, a two-month delay to the end restrictions was considered as part of the scenario analysis. Regarding vaccine uptake, at the time of the study this stood at 90% within the 41% of the BNSSG population that had been offered vaccination up to that point. Local expectations of ultimate vaccine uptake centred around 90%. However, there was uncertainty around whether the hitherto strong uptake rates would be replicated in the younger age groups due to be offered vaccination next. As such, 75% ultimate uptake was considered as part of the scenario analysis. Finally, the aforementioned lengths of stay at the time of the study were higher than those defined as key performance indicators by senior community service provider managers. Accordingly, their target 10, 21 and 28 day mean lengths of stay for P1-3 respectively were included in the scenario analysis.

### Demand projection

In order to project intermediate care demand directly caused by COVID-19 infection, an epidemiological model of the ‘Susceptible-Exposed-Infectious-Recovered’ type was used [24]. Developed in the BNSSG healthcare system, this had been in routine use for estimating the future number of COVID-19 patients requiring acute and intermediate care services. Each week the model was recalibrated with the latest data and scenario assumptions, including assessments of future vaccine uptake and public mobility (the latter being estimated by the level of societal restriction currently or expected to be in place). Results were used for a variety of operational purposes, such as informing the necessity of opening new COVID-19 wards. For this study, the model was calibrated using the four combinations of vaccination uptake and societal restrictions as specified in Table 1.

For projections of intermediate care demand not directly caused by COVID-19 infection, a regression model linking non-COVID-19 emergency admissions to levels of public mobility was used (Figure 2). Previous research conducted within the BNSSG system had identified evidence of a moderately strong association between these two variables, with an R-squared of 0.62 [25]. The general premise was that with higher levels of mobility there is increased risk of the kind of events that generate such demand, e.g. sports injuries and road traffic accidents. For this study, the regression model was calibrated using the two combinations of societal restrictions as specified in Table 1.

**Figure 2.**
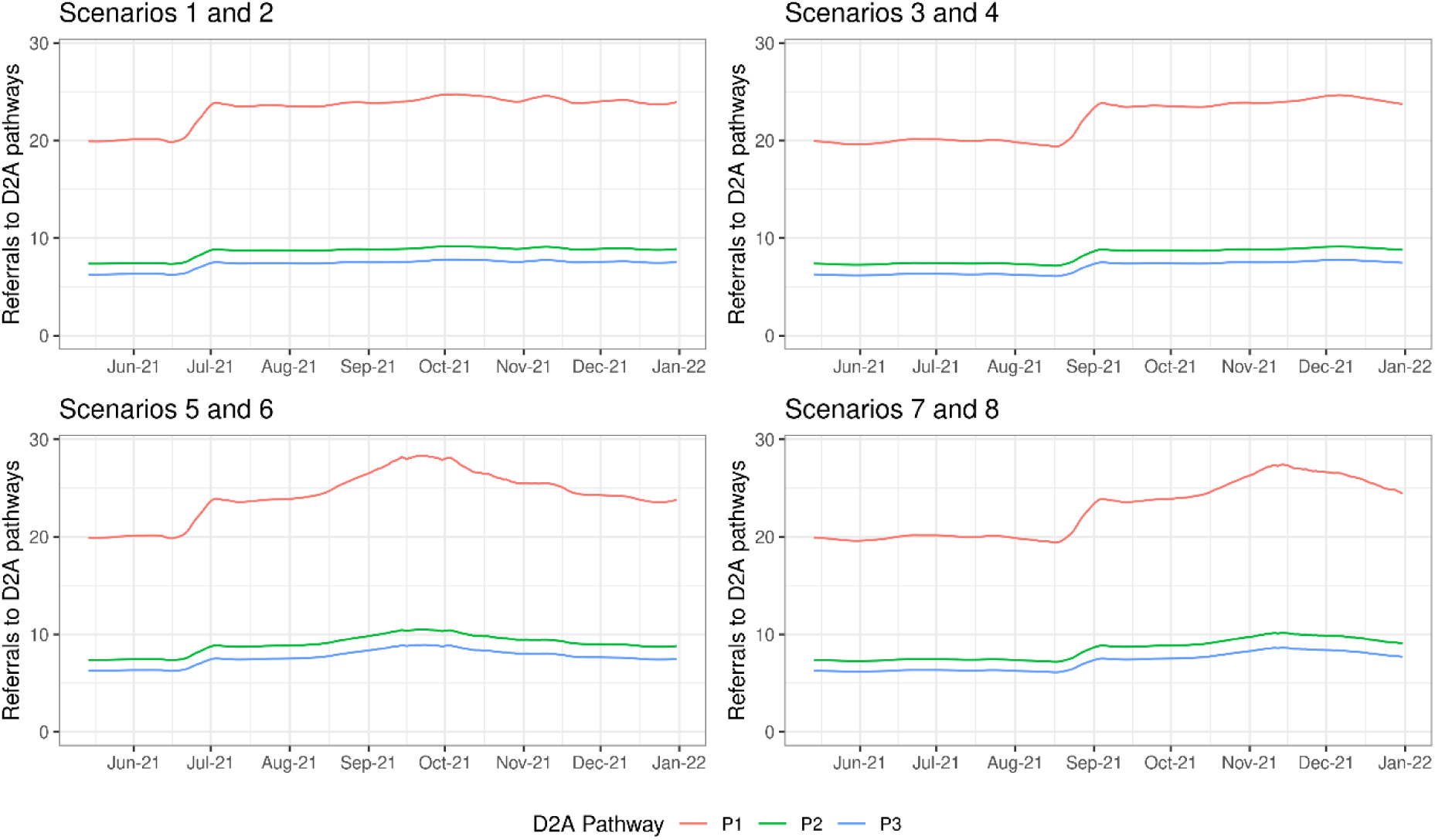
Projections of demand for intermediate care Discharge to Assess (D2A) Pathways P1-3, for each of the considered scenarios.

Both the above demand models produced acute admission projections which were converted to D2A pathway demand through the following assumptions: a 6-day mean acute care length of stay; a 19% proportion of acute discharges requiring intermediate care; and a 54%, 20% and 17% split of intermediate care patients to D2A pathways P1-3 respectively (the remaining 9% are accounted for by specialist end-of-life pathways).These values were estimated using data from the BNSSG system at the time of the study.

Combining both the COVID-19 and non-COVID-19 components of projected demand yielded the total demand expected under the various scenarios (Figure 2). From this, the fairly immediate effects of relaxing societal restrictions on non-COVID-19 demand is evident (see the step change in late June for Scenarios 1, 2, 5, 6, and in late August for Scenarios 3, 4, 7, 8). The easing of societal restrictions also impacts upon COVID-19 demand but, due to disease transmission dynamics, there is a time lag before this peaks. In fact, with 90% vaccine uptake, there is negligible transmission and so negligible demand (Scenarios 1-4). With 75% uptake, however, a third wave is forecast in autumn 2021 (Scenarios 5-8), noting that a similar timing had been estimated in other epidemiological studies at that time [26].

### Computer simulation modelling

Computer simulation is considered a well-established and conceptually-appropriate approach to modelling demand and capacity in healthcare systems [13,14]. In this study, ‘discrete time’ simulation was used to dynamically model the flow of individual patients from acute discharge readiness (i.e. to become an ‘arrival’ at the start of a D2A waiting list) through to completion of intermediate care. Essentially this involves simulating the arrival of simulated individual patients (see *Method; Demand projection*), commencement of intermediate care in the relevant D2A pathway (for which the patient may have to wait, depending on available capacity), and their departure from the service (determined by their length of stay).

A separate model was constructed for each of the P1-3 D2A pathways and the eight scenarios considered (Table 1). At the start of each simulation, the initial occupancy and waiting list size was set equal to their actual values as of 14 May 2021. Within each simulation, each future day (the ‘discrete time’ interval used in this study) was simulated consecutively, with instances of the above-mentioned three events performed in line with the simulation schedule (the arrivals onto the D2A pathway which were due to occur that day; how many patients were due to complete their D2A pathway that day; and the commencement of intermediate care provided there were patients waiting for care and available capacity). Each simulation was run until 31 December 2021, with 200 replications performed for each simulation in order to capture the realistic effect of variability (with respect to arrivals and lengths of stay). Results for each simulation were calculated from the outputs of these replications.

Simulations were run over a range of intermediate care capacity configurations, in order to reflect various choices or targets considered by healthcare managers and planners in the BNSSG health system. For each of these, the key output measure of interest was the total cost of both intermediate care service provision (calculated from the modelled capacity levels) and the acute capacity required to support any delays to discharge (calculated from the mean number of acute beds blocked). In calculating this, the unit costs (per patient per day) used are £346 for acute and £125 for P1 [27], £150 for P2 [28], and £164 for P3 (local data). The respective ratio of unit costs is therefore approximately 14:5:6:7. All models and analyses were produced in the R software environment for statistical computing. Full details on the modelling method and calibration are provided in Supplementary Material A.

## Results

Under the *Baseline* scenario, the estimated cost-optimal capacity requirements for D2A Pathways P1-3 are 700, 275 and 335 respectively (Scenario 1; upper panels Figure 3 and Table 2). These are significantly higher than the capacities at the time of the study (546, 177 and 172 respectively). This is partly explained by the forecast increased demand for D2A services (due to relaxations in COVID-19 related restrictions – see Figure 2), but is also due to a more cost-efficient resource allocation; in which high-cost acute discharge delays (lower panels Figure 3 and Table 2) are reduced from their levels at the time of the study (64, 62 and 83 respectively) through a lower D2A service occupancy (middle panels Figure 3 and Table 2, c.f. 100%, 85% and 100% at the time of the study) brought about by higher D2A service capacity. It is these lower discharge delays that dictate the cost-optimal capacity requirement – this is evident from the asymmetry in the cost curves (upper panels Figure 3) with the large cost increases at capacities below the cost-optimal driven by acute care queueing (upper panels c.f. lower panels Figure 3) and the relatively lower cost increases at above-optimal capacities driven by the D2A service provision cost (which increases linearly with capacity).

**Table 2.** Estimated cost-optimal capacities (daily maximum number of visits for P1; beds for P2 and P3) and corresponding summarised results for mean (95% CI) D2A service occupancy (expressed as a percentage of total capacity) and mean (95% CI) number of acute beds blocked. Note, these results correspond to those illustrated by the dashed vertical lines in Figure 4.

| Scenario | Cost-optimal capacity |  |  | D2A service occupancy %, mean (95% CI) |  |  | Acute beds blocked, mean (95% CI) |  |  |
| --- | --- | --- | --- | --- | --- | --- | --- | --- | --- |
|  | P1 | P2 | P3 | P1 | P2 | P3 | P1 | P2 | P3 |
| 1 | 700 | 275 | 335 | 89 (70-91) | 88 (77-96) | 88 (80-93) | 0.88 (0-12) | 2.68 (0-19) | 6.89 (0-30) |
| 2 | 550 | 200 | 220 | 88 (69-100) | 89 (77-96) | 91 (79-97) | 0.65 (0-10) | 1.70 (0-14) | 4.02 (0-25) |
| 3 | 680 | 260 | 320 | 87 (70-100) | 87 (77-94) | 87 (79-93) | 1.48 (0-19) | 5.04 (0-22) | 6.91 (0-25) |
| 4 | 540 | 190 | 210 | 85 (63-100) | 88 (76-96) | 89 (79-97) | 0.77 (0-11) | 3.52 (0-17) | 4.93 (0-22) |
| 5 | 780 | 295 | 360 | 84 (63-100) | 86 (76-92) | 86 (78-91) | 0.96 (0-15) | 3.92 (0-22) | 7.24 (0-28) |
| 6 | 610 | 215 | 240 | 83 (62-100) | 86 (75-94) | 87 (76-94) | 0.61 (0-9) | 2.80 (0-17) | 3.17 (0-17) |
| 7 | 750 | 275 | 330 | 82 (63-100) | 85 (75-91) | 86 (79-91) | 1.11 (0-16) | 6.17 (0-22) | 10.9 (0-26) |
| 8 | 580 | 200 | 220 | 81 (62-100) | 86 (75-93) | 87 (78-94) | 0.99 (0-14) | 5.48 (0-19) | 7.17 (0-21) |

**Figure 3.**
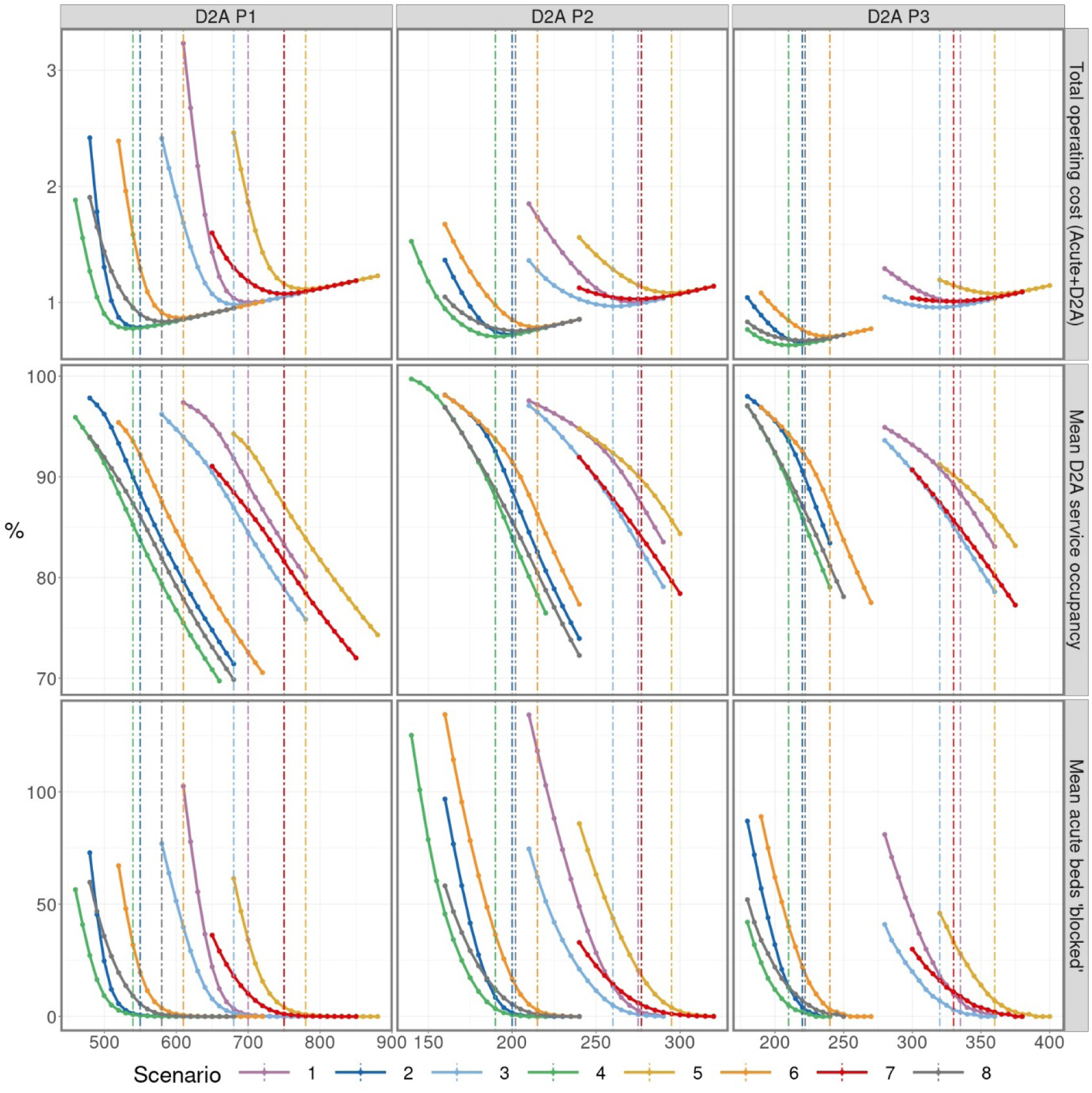
Simulated results for total operating cost of D2A service and acute delays (indexed on baseline); mean D2A service occupancy, expressed as a percentage of total capacity; and mean number of acute beds ‘blocked’ (i.e. acute patients whose discharge is delayed due to insufficient D2A service capacity). Simulations were performed over the period 14 May 2021 to 31 December 2021 for each of the three D2A pathways for a range of considered service capacities and scenarios (Table 1). Dashed vertical lines highlight the results corresponding to the cost-optimal capacity, as determined by the lowest total weekly cost.

The particular level of acute discharge delays deemed optimal is dependent on the relative acute care and D2A unit costs: the greater the former are in relation to the latter, then the more acute bed blocking is penalised at the expense of greater D2A under-utilisation (i.e. lower D2A occupancy). An example of this is the higher number of acute beds blocked accepted for P3 (mean 6.89) when compared to P1 (0.88) and P2 (2.68), given the 14:5:6:7 ratio between acute and P1-3 unit costs. Variability in acute discharge delays, alongside D2A service occupancy, is illustrated in Figure 4. Such information can be useful alongside average measures, particularly for service planning (e.g. in order to gauge potential workforce requirements for managing high D2A occupancy, or whether or not to postpone elective treatments because of significant acute discharge delays).

**Figure 4.**
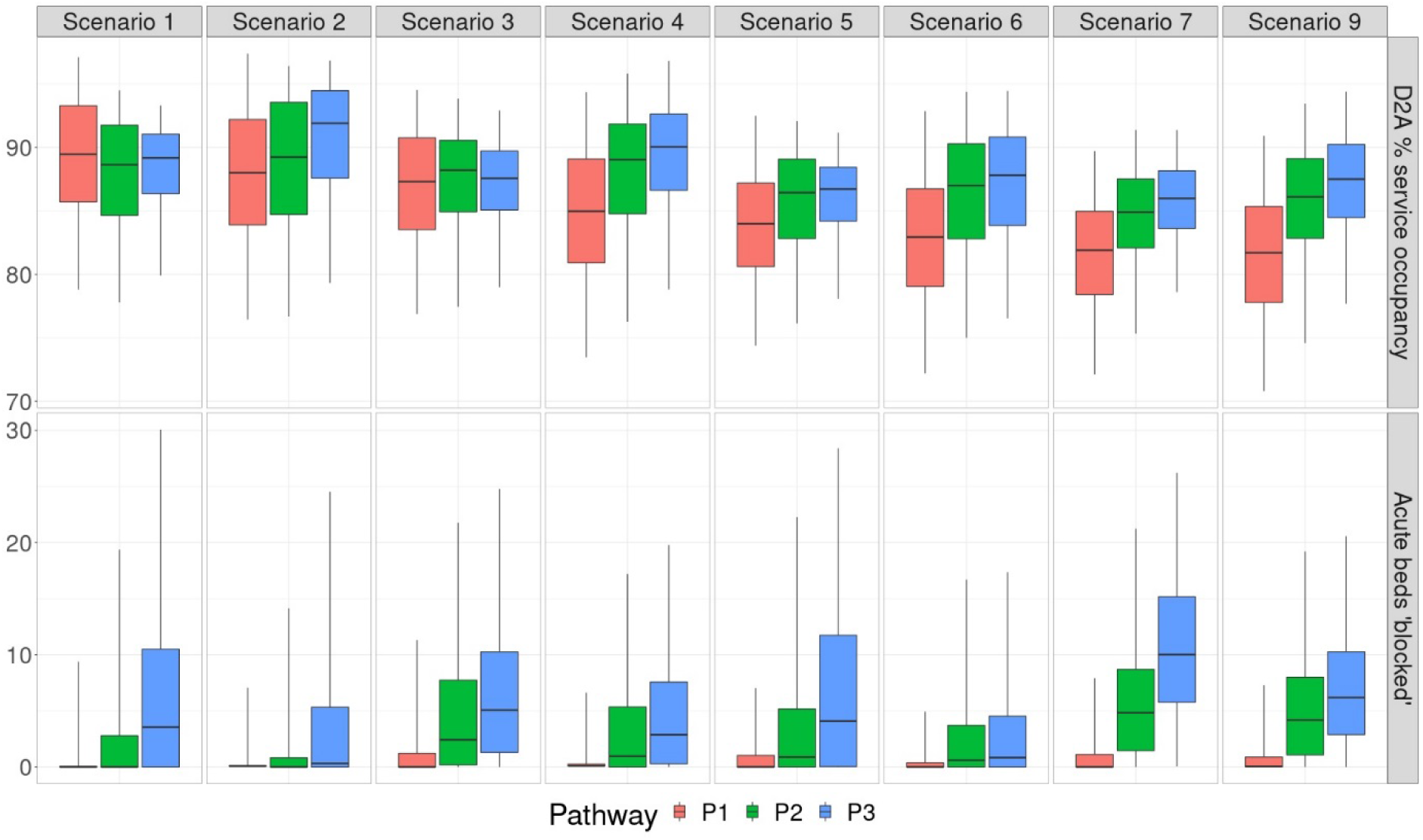
Box-whisker plots illustrating, for the cost-optimal D2A capacities, the median and interquartile range of D2A service occupancy (expressed as a proportion of total capacity) and number of acute beds blocked.

Greater hospital demand, resulting from a reduction in vaccination uptake in the baseline assumption of 90% to 75%, is shown to increase cost-optimal D2A capacity requirement by between 7% and 11% ceteris paribus (Scenario 5 c.f. Scenario 1). Even with this additional capacity, and a 2-6% lower occupancy, the additional demand for D2A services leads to a 16% increase in mean acute discharge delays across the D2A pathways, with total costs rising by up to 11.4%. Additionally, if the end to societal restrictions is delayed by two months, then this decreases the number of hospital admissions (both directly and indirectly related to COVID-19) occurring within the remainder of 2021, leading to a reduction in the cost-optimal capacity requirement for the year (Scenario 7 c.f. Scenario 5). If, at the same time, length of stay can be reduced from those actually occurring at the time of the study to target levels, then the cost-optimal capacity requirement can be significantly reduced to levels similar to those at the time of the study (i.e. 580, 200 and 220 c.f. the 546, 177 and 172 as of May 2021). Thus the effect of the greater demand for D2A services can be mitigated while at the same time attaining a more cost-efficient service with reduced acute discharge delays (meaning less wastage and improved patient experience).

Supplementary Material B provides a sensitivity analysis with respect to the assumed 19% for acute admissions requiring D2A support upon discharge. While this figure is empirically-derived, its validity over the forecasted period may be affected by an uncertain future patient casemix. The sensitivity analysis confirms that increasing (30% complex discharges) and decreasing (10% complex discharges) arrival rates impacts both the total system costs and the cost-optimal capacities. Due to ‘economies of scale’, the mean cost per delayed discharge reduces slightly as output increases. Supplementary Material C contains simulated results with no limit on D2A capacity, providing an assessment of unconstrained D2A occupancy over the forecasted period. Supplementary Material D contains the appropriate research checklist used to ensure unambiguous reporting of our study.

## Discussion

### Main findings

While there has been speculation that system-level costs can be reduced by greater downstream investment in complex care pathways, there has been a deficit of clear evidence to support these anecdotal claims. Through development and application of a conceptually-appropriate computer model to a large health economy operating the D2A model of care used in the UK, our study demonstrates the potential value of increasing intermediate care capacity, specifically during recovery from the COVID-19 pandemic. Results show that the additional service provision cost is offset by savings associated with reduced delays to acute discharge, thus suggesting that balancing capacity across the complex care pathway is not a zero-sum game, and that net costs can be reduced through appropriate capacity allocation. Alongside financial advantages, further benefits of lower acute discharge delays – while not quantitatively assessed – are likely to extend to improved patient outcomes and experience through reduced medical complications and impacts on patient activation and independence.

A particular finding of this study has been the significantly greater cost increases for equivalent amounts of capacity under the cost-optimal level than above it (i.e. the ‘asymmetry’ noted in *Results*). This is explained by the cost of intermediate care provision increasing linearly with capacity, whereas acute discharge delay costs increase rapidly and nonlinearly below a certain level of capacity (Figure 3, upper panels). This study also finds that pursuit of zero acute discharge delay is unlikely to result in a cost-optimal service. To eliminate acute discharge delays would require uneconomically high levels of intermediate care capacity, substantial amounts of which would be unused some of the time (Figure 3, lower panels c.f. middle panels). While the precise extent of this is dependent on the inherent variability of the pathway (greater slack in occupancy is usually always required with more variable patient arrivals and length of stay [29]), this study finds that some amount of acute discharge delay is likely to be economically essential, and provides readers with a description of an approach to its calculation alongside the means to do so, i.e. the open-source software.

### Practical implications

Taking the main findings in turn; increasing intermediate care capacity is a challenging prospect when operating in a financially constrained environment and the health economy of this study, like many others in the UK, is running at a deficit. However, the cost must be viewed against the (greater) acute care cost saving. Yet, as ‘date ready for discharge’ is not always recorded in acute hospital datasets, it is difficult to quantify the magnitude of this cost and thus set out the scope for potential savings in making any case for change. Some scale is, however, attainable from one relevant identified study which estimated that the national cost “*of treating older patients in hospital who no longer need to receive acute clinical care*” was approximately £820m [30].

The next main finding is that there are higher costs where intermediate care capacities are under the cost-optimal level than above it. Thus, from a practical standpoint, if there is doubt about the value of this level, it is better to pitch higher than lower in setting the intermediate care capacity requirement. Finally, given the economic need for at least some acute discharge delays, this study cautions any use of performance targets to completely eliminate acute bed blockages; instead, hospital managers should aim for an amount which reflects wider pathway performance. This may pose a challenge for health systems with insufficient integration of constituent organisations (both in the financial and political senses).

For the healthcare system considered in this study, results were shared with senior managers on 2 June 2021. This was less than three weeks following the most recent input data (14 May 2021), with modelling and analysis performed rapidly in order to ensure relevancy. It should be acknowledged, and expected, that, given the paucity of past modelling in this domain (both within our system and more widely), a certain level of familiarity needs to be gained in crystallising practical understanding and actions. Combined with the pressures of COVID-19, this reduced the ability to associate particular decisions with model outputs in the short term. However, in the longer term, the modelled results (specifically the ‘baseline’ Scenario 1) have played a central role in a Business Case (written in summer 2021) setting out increased D2A capacity requirements for the remainder of the year.

### Limitations

As a simplifying assumption, the computer model works on the basis of a ‘7 day’ operational service, with no effect of day of week on patient admission and discharge (this is not always the case as there can be some delay in transferring patients at weekends). The effect of this is to underestimate acute discharge delays, with the actual number of acute beds required being greater than that modelled (i.e. in Figure 3, lower panels). This is important since insufficient acute bed capacity can propagate upstream to cause blockages in the emergency department as well as prompting elective surgery cancellations.

Another aspect of the model requiring some attention is the assumption that capacity is fixed across the modelled period. With expected fluctuations in demand (Figure 2), an appropriately-varying capacity may appear reasonable. However, senior managers had stipulated that just a single capacity value was required, given the intended strategic long-term use of the model and an appreciation that any capacity shortfalls would be responded to on a more operational basis, e.g. with temporary procurement of additional capacity from the independent sector.

In terms of model validation, an objective assessment was not possible since the Business Case had not been implemented at the time of writing, and a period of time would be required in order to robustly assess the reliability of model outputs. In general terms, validity was promoted through use of a model that was calibrated using real-life data (thus obviating the need for unsupported assumptions) and, as well as being conceptually appropriate, it was straightforward to implement (thus avoiding the potential pitfalls associated with overly complex methods, such as over-fitting or over-specification). Additional confidence was gained through having worked closely alongside NHS analysts, managers and clinicians throughout the project, thus increasing scrutiny at each step of the modelling journey. While not a comprehensive assessment of output validity, some attempts can be made. First, the mean D2A service occupancy (Table 2) is broadly within the 80-90% expected for large capacity pools [30]. Second, observations since 14 May 2021 are approximately consistent with expectations given what has since transpired: an ultimate vaccination uptake of 83%, a one-month delay to the ending of societal restrictions, and D2A lengths of stay unmoved from their baseline values, i.e. behaviour between Scenarios 1 and 3 (see Table 2). While higher D2A capacity is proposed in the Business Case, in the three months since 14 May 2021 it remained fairly unchanged at 552 (P1), 153 (P2) and 171 (P3). From Figure 3 (Scenarios 1 and 3), these capacity levels would suggest very large numbers of acute beds blocked – which is corroborated given mean figures of 71 (P1), 57 (P2) and 57 (P3) in the three months since 14 May 2021.

### Further research

There is a plethora of directions for future work. First, investigators may seek to quantify the impact of acute discharge delay on patient outcomes and experience, and thus develop a multi-objective analysis not fixated solely on cost. This would be expected to further penalise acute discharge delays, leading to a higher intermediate care capacity requirement.

Second, in addition to considering ‘step-down’ care from acute to community care, the model could be extended to include ‘step up’ care (i.e. where community care is provided proactively to patients in their usual residence in order to prevent an emergency admission). Given that community service providers have a fixed resource for addressing both such aspects of care, modelling could be useful in calculating the optimal demarcation.

Third, the model could be extended further downstream into social care, as insufficient capacity here causes similar issues for intermediate care that intermediate care, in turn, causes for acute hospitals. With the recently published White Paper on the future of the NHS paving the way for enhanced health and care integration, healthcare systems may soon be forced to do more to confront such matters, with modelling serving as a potentially valuable asset.

## Data Availability

Model code can be found at https://github.com/nhs-bnssg-analytics/ipacs-v1-model.

## Data and material

Model code can be found at https://github.com/nhs-bnssg-analytics/ipacs-v1-model.

## Acknowledgements

The authors gratefully acknowledge the support of Rosie Hume and Julie Kell.

## References

[1] El Bcheraoui C, Weishaar H, Pozo-Martin F, Hanefeld J. Assessing COVID-19 through the lens of health systems’ preparedness: time for a change. Globalization and Health. 2020 Dec;16(1):1–5. https://doi.org/10.1186/s12992-020-00645-5.

[2] McCabe R, Schmit N, Christen P, D’Aeth JC, Løchen A, Rizmie D, Nayagam S, Miraldo M, Aylin P, Bottle A, Perez-Guzman PN. Adapting hospital capacity to meet changing demands during the COVID-19 pandemic. BMC medicine. 2020 Dec;18(1):1–2. https://doi.org/10.1186/s12916-020-01781-w.

[3] Verhagen MD, Brazel DM, Dowd JB, Kashnitsky I, Mills MC. Forecasting spatial, socioeconomic and demographic variation in COVID-19 health care demand in England and Wales. BMC medicine. 2020 Dec;18(1):1–1. https://doi.org/10.1186/s12916-020-01646-2.

[4] Wood RM, McWilliams CJ, Thomas MJ, Bourdeaux CP, Vasilakis C. COVID-19 scenario modelling for the mitigation of capacity-dependent deaths in intensive care. Health care management science. 2020 Sep;23(3):315–24. https://doi.org/10.1007/s10729-020-09511-7.

[5] Li M, Vanberkel P, Carter AJ. A review on ambulance offload delay literature. Health care management science. 2019 Dec;22(4):658–75. https://doi.org/10.1007/s10729-018-9450-x.

[6] Micallef A, Buttigieg S, Tomaselli G, Garg L. Defining delayed discharges of inpatients and their impact in acute hospital care: a scoping review. International Journal of Health Policy and Management. 2020 Jun 29. https://doi.org/10.34172/IJHPM.2020.94.

[7] Everall AC, Guilcher SJ, Cadel L, Asif M, Li J, Kuluski K. Patient and caregiver experience with delayed discharge from a hospital setting: a scoping review. Health Expectations. 2019 Oct;22(5):863–73. https://doi.org/10.1111/hex.12916.

[8] Tenforde MW, Kim SS, Lindsell CJ, Rose EB, Shapiro NI, Files DC, Gibbs KW, Erickson HL, Steingrub JS, Smithline HA, Gong MN. Symptom duration and risk factors for delayed return to usual health among outpatients with COVID-19 in a multistate health care systems network—United States, March–June 2020. Morbidity and Mortality Weekly Report. 2020 Jul 31;69(30):993. https://doi.org/10.15585/mmwr.mm6930e1.

[9] Luceri F, Morelli I, Accetta R, Mangiavini L, Maffulli N, Peretti GM. Italy and COVID-19: the changing patient flow in an orthopedic trauma center emergency department. https://doi.org/10.1186/s13018-020-01816-1.

[10] Litvak E, Keshavjee S, Gewertz BL, Fineberg HV. How Hospitals can Save Lives and Themselves: Lessons on Patient Flow from the COVID-19 Pandemic. Annals of Surgery. 2021 Jul 1;274(1):37–9. https://doi.org/10.1097/SLA.0000000000004871.

[11] Sezgin D, O’Caoimh R, O’Donovan MR, Salem MA, Kennelly S, Samaniego LL, Carda CA, Rodriguez-Acuña R, Inzitari M, Hammar T, Holditch C. Defining the characteristics of intermediate care models including transitional care: an international Delphi study. Aging clinical and experimental research. 2020 Nov;32(11):2399–410. https://doi.org/10.1007/s40520-020-01579-z.

[12] NHS. 2021. Hospital Discharge and Community Support: Policy and Operating Model. https://assets.publishing.service.gov.uk/government/uploads/system/uploads/attachment_data/file/999443/hospital-discharge-and-community-support-policy-and-operating-model.pdf.

[13] Brailsford SC, Harper PR, Patel B, Pitt M. An analysis of the academic literature on simulation and modelling in health care. Journal of simulation. 2009 Sep 1;3(3):130–40. https://doi.org/10.1057/jos.2009.10.

[14] Fone D, Hollinghurst S, Temple M, Round A, Lester N, Weightman A, Roberts K, Coyle E, Bevan G, Palmer S. Systematic review of the use and value of computer simulation modelling in population health and health care delivery. Journal of Public Health. 2003 Dec 1;25(4):325–35. https://doi.org/10.1093/pubmed/fdg075.

[15] Gupta D, Denton B. Appointment scheduling in health care: Challenges and opportunities. IIE transactions. 2008 Jul 21;40(9):800–19. https://doi.org/10.1080/07408170802165880.

[16] Chan CW, Farias VF, Bambos N, Escobar GJ. Optimizing intensive care unit discharge decisions with patient readmissions. Operations research. 2012 Dec;60(6):1323–41. https://doi.org/10.1287/opre.1120.1105.

[17] Patrick J, Nelson K, Lane D. A simulation model for capacity planning in community care. Journal of Simulation. 2015 May 1;9(2):111–20. https://doi.org/10.1057/jos.2014.23.

[18] Demirbilek M, Branke J, Strauss A. Dynamically accepting and scheduling patients for home healthcare. Health care management science. 2019 Mar;22(1):140–55. https://doi.org/10.1007/s10729-017-9428-0.

[19] Palmer R, Fulop NJ, Utley M. A systematic literature review of operational research methods for modelling patient flow and outcomes within community healthcare and other settings. Health Systems. 2017 Mar 23:1–21. https://doi.org/10.1057/s41306-017-0024-9.

[20] Monks T, Worthington D, Allen M, Pitt M, Stein K, James MA. A modelling tool for capacity planning in acute and community stroke services. BMC health services research. 2016 Dec;16(1):1–8. https://doi.org/10.1186/s12913-016-1789-4.

[21] Wood RM, Murch BJ. Modelling capacity along a patient pathway with delays to transfer and discharge. Journal of the Operational Research Society. 2020 Oct 2;71(10):1530–44. https://doi.org/10.1080/01605682.2019.1609885.

[22] UK Government. 2021. COVID-19 Response – Spring 2021 (Summary): Roadmap out of lockdown. Available at: https://www.gov.uk/government/publications/covid-19-response-spring-2021/covid-19-response-spring-2021-summary.

[23] Mahase E. Covid-19: Is it safe to lift all restrictions in England from 21 June?. https://doi.org/10.1136/bmj.n1399.

[24] Booton RD, MacGregor L, Vass L, Looker KJ, Hyams C, Bright PD, Harding I, Lazarus R, Hamilton F, Lawson D, Danon L. Estimating the COVID-19 epidemic trajectory and hospital capacity requirements in South West England: a mathematical modelling framework. BMJ open. 2021 Jan 1;11(1):e041536. http://dx.doi.org/10.1136/bmjopen-2020-041536.

[25] Powell AL, Wood RM. Projecting the effect of easing societal restrictions on non-COVID-19 emergency demand in the UK: Statistical inference using public mobility data. The International Journal of Health Planning and Management. 2021 Sep;36(5):1936–42. https://doi.org/10.1002/hpm.3265.

[26] Iacobucci G. Covid-19:”Freedom day” in England could lead to “significant third wave of hospitalisations and deaths,” modelling predicts. https://doi.org/10.1136/bmj.n1789.

[27] NHS Improvement. 2019. 2017/18 Reference Costs. https://webarchive.nationalarchives.gov.uk/ukgwa/20200501111106/ https://improvement.nhs.uk/resources/reference-costs/.

[28] NAIC. 2017. National Audit of Intermediate Care: Summary Report England 2017. https://s3.eu-west-2.amazonaws.com/nhsbn-static/NAIC+(Providers)/2017/NAIC+England+Summary+Report+-+upload+2.pdf.

[29] Pratt AC, Wood RM. Addressing overestimation and insensitivity in the 85% target for average bed occupancy. International Journal for Quality in Health Care. 2021;33(3):mzab100. https://doi.org/10.1093/intqhc/mzab100.

[30] National Audit Office. 2016. “Discharging Older Patients from Hospital”. https://www.nao.org.uk/report/discharging-older-patients-from-hospital/.

