## Supplementary material for "Optimising the balance of acute and intermediate care capacity for the complex discharge pathway: computer modelling study during COVID-19 recovery in England"

**Supplementary Material A: Pathway simulation model**

The pathway computer simulation model developed for the purposes of this study aims at determining required capacity for the three D2A step-down intermediate care pathways independently (P1, P2 and P3). The model was coded in R (version 3.6.4) as an initial improvement to a Microsoft Excel based deterministic model used for capacity planning by the collaborating health and care organisations. All simulation parameters were estimated in collaboration with the healthcare services involved. Figure 1 (main paper) illustrates the general flow of the patients simulated in this model from arrival and stay in hospital, to discharge into P1-3 pathways.

From a modelling perspective, there are two types of pathways depending on the type of service patients require and the related capacity involved: visit-based and bed-based. Patients entering a visit-based care pathway (P1) reside in their usual place of residence and community care is provided by one or more care workers via daily visits. For this type of pathway, capacity is based on the number of (time) slots and the required number of visits per slot. Patients needing bed-based care (P2 and P3) will transfer to a step-down facility after discharge from the acute hospital for a pre-determined length of stay, hence the capacity is evaluated in terms of the number of community health beds required.

Costs used in the model are presented in Table SM.A.2. As the focus of this paper is the identification of cost-optimal intermediate care capacity through minimising system costs, we chose to use relative, rather than absolute, costs. Plots 3 (main paper), SM.B.1, SM.B.2 and SM.B.3 display all costs indexed to the minimum ('cost-optimum') of the baseline scenario (Scenario 1) for each pathway.

***The visit-based care pathway model***

Daily referral rates into P1 were generated using a Poisson process where the mean for each day has been estimated using the prediction models described in subsection 'Demand projection' of the main paper. Patients who are referred but unable to be discharged into P1 remain in hospital and represent a delayed discharge. Reflecting current data, the duration of P1 service is sampled from a normal distribution. The initial daily visit requirements, and the final daily visit requirements are sampled from normal distributions. The mean and standard deviation of the baseline duration of service were estimated from up-to-date data provided by intermediate care providers. Although, the number of visits required per patient-day is not recorded on a regular basis, the intermediate care service base their planning on an average of three care visits per day. In practice, it is known that the number of visits tapers over the duration of service. Also, patients in P1 may initially require two care staff per visit. Accordingly, we set the mean of initial number of visits required to four (truncated to an upper limit of six) and the mean of the final number of visits to two. In the model, a sequence of visits from the sampled initial number to the sampled final number of visits across a duration of service was generated per simulated patient. For example:

- If the initial number of visits is sampled as six, and
- the final number of visits is sampled as two, and
- the duration of service is sampled as 10 days,
- then, the visit sequence for the patient is [6,6,5,5,4,4,3,3,2,2].

The capacity is the number of visits in the system, i.e., the number of patients admitted into P1 ('slots'), multiplied by the average number of visits per day. Patients with a long duration of service, and/or a high initial/end visit requirement subsequently may prevent patients with lower service requirements from entering the P1 system. For this reason, if there are no available resources to start service for a new patient immediately, an arriving patient is scheduled to start on the following day. A patient whose visit sequence can be integrated into the available P1 capacity will be scheduled immediately.

#### *The bed-based care pathway model*

As in the visit-based model, daily referrals were generated using a Poisson distribution with daily means estimated from the demand prediction models described in in subsection 'Demand projection' of the main paper. For each patient, the length of stay was sampled from a lognormal distribution with parameters estimated using the data obtained from the CCG. Any patient who is referred but unable to be transferred into the assigned bedded pathway will remain in hospital as a delayed discharge. In this model, the capacity is represented by the number of beds available and any patient who enters the corresponding pathway occupies the bed until the end of their length of stay. The maximum capacity needed is represented by the number of beds required to have zero delayed discharges from the acute hospital. To reflect the current situation in the considered system each bed-based care pathway, in this case P2 and P3, is treated independently.

The input parameter values used in the pathway simulation model are presented in Table SM.A.1.

**Table SM.A.1.** Pathway simulation model input parameters (LoS: Length of Stay). * Estimated from care provider data.

| Pathway | Number of simulation runs (replications) | Initial occupancy on 14 May 2021* | Mean LoS* | LoS Distribution | Mean Arrival Rate | Arrivals Distribution | Proportion of patients entering pathway (%) * |
|---|---|---|---|---|---|---|---|
| P1 | 200 | 184 | 13 | Normal | Daily projection | Poisson | 54 |
| P2 | 200 | 151 | 29 | Lognormal | Daily projection | Poisson | 20 |
| P3 | 200 | 172 | 43 | Lognormal | Daily projection | Poisson | 17 |

The cost data used to simulate system costs are presented in Table SM.A.2.

**Table SM.A.2.** Simulation model cost parameters, cost ratios, and sources.

| | Average cost of weekly service | Relative Cost ratios | Source of costs |
|---|---|---|---|
| P1 | £875 | 5 | 2017/18 NHS reference costs |
| P2 | £1,050 | 6 | National Audit of Intermediate Care 2017/18 |
| P3 | £1,150 | 7 | System costs from Bristol care system |
| Acute | £2422 | 14 | 2017/18 NHS reference costs |

To determine the optimum capacity which minimizes the total cost of acute delayed discharges and the cost of providing surplus capacity, the simulation model calculated the overall cost across a range of feasible capacities. The capacity is either the number of visits or the number of beds available to the system. To determine the lower and upper bound of the range of capacity required, the simulation was first run assuming infinite available capacity to find the capacity needed for zero acute hospital delayed discharges. The upper bound was set to 75% quantile of the maximum capacity needed. A lower bound that reflects the evolution of the costs with respect to capacity was set by visual inspection of the outputs. The total daily cost we considered includes the delayed discharge cost in the acute and the service cost in intermediate care. The average cost of delayed discharge in acute care is calculated as the cost of one hospital day multiplied by the average delay per simulated patient and the number of patients discharged per week.

**Supplementary Material B: Sensitivity analysis accounting for uncertainty regarding age and multiborbidity of acute admissions**

In our baseline model we assumed that 19% of acute discharges are complex and patients enter one of the three community care pathways after discharge. However, we recognise that this figure may vary based on different patient-related characteristics such as age, presence of chronic conditions and/or comorbidities. In order to account for this uncertainty, we present a sensitivity analysis over the proportion of complex discharges by running simulation assuming a complex discharge proportion of 10% (Figures SM.B.1 and SM.B.2) and 30% (Figures SM.B.3 and SM.B.4).

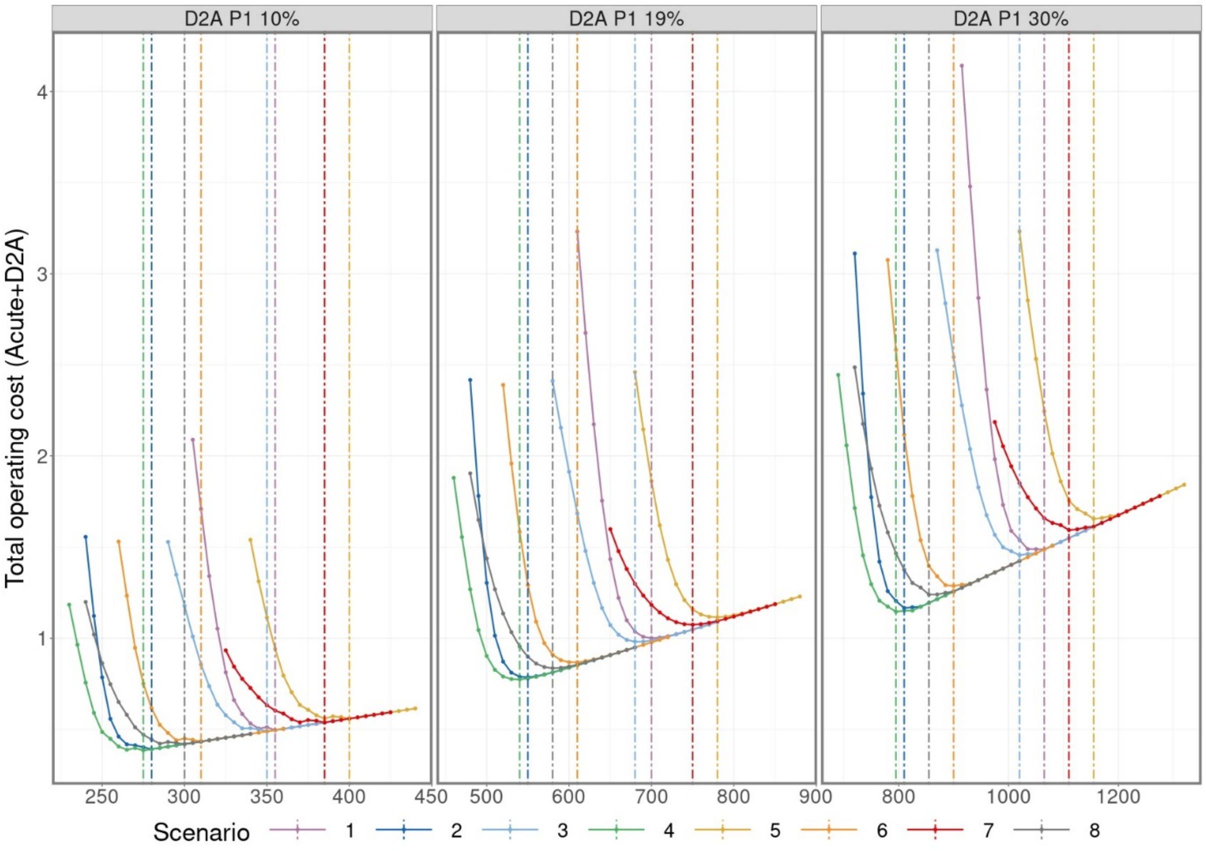


**Figure SM.B.1.** Weekly cost of acute delay and pathway care community service for capacity configurations for Pathway 1 with 10%, 19% (baseline) and 30% of acute admissions requiring complex community services at discharge.

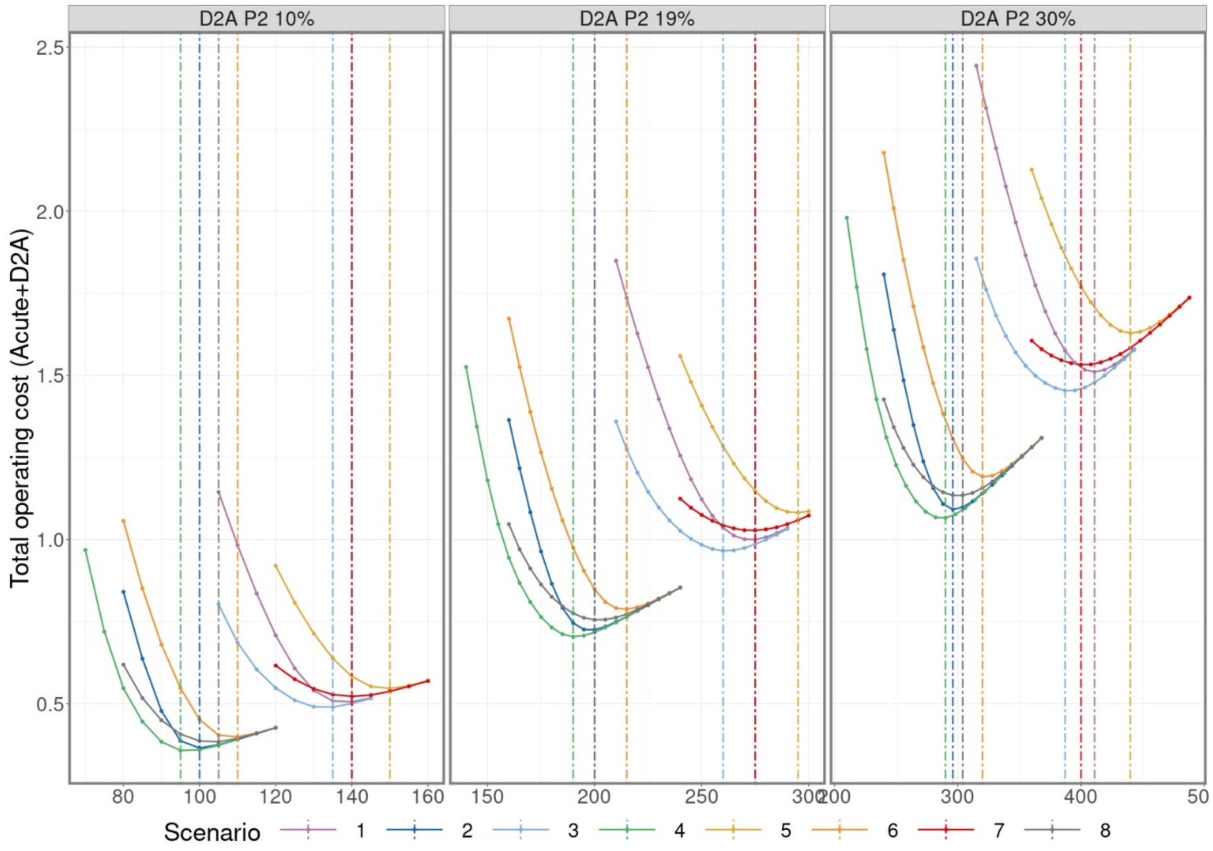


**Figure SM.B.2.** Weekly cost of acute delay and pathway care community service for capacity configurations for Pathway 2 with 10%, 19% (baseline) and 30% of acute admissions requiring complex community services at discharge.

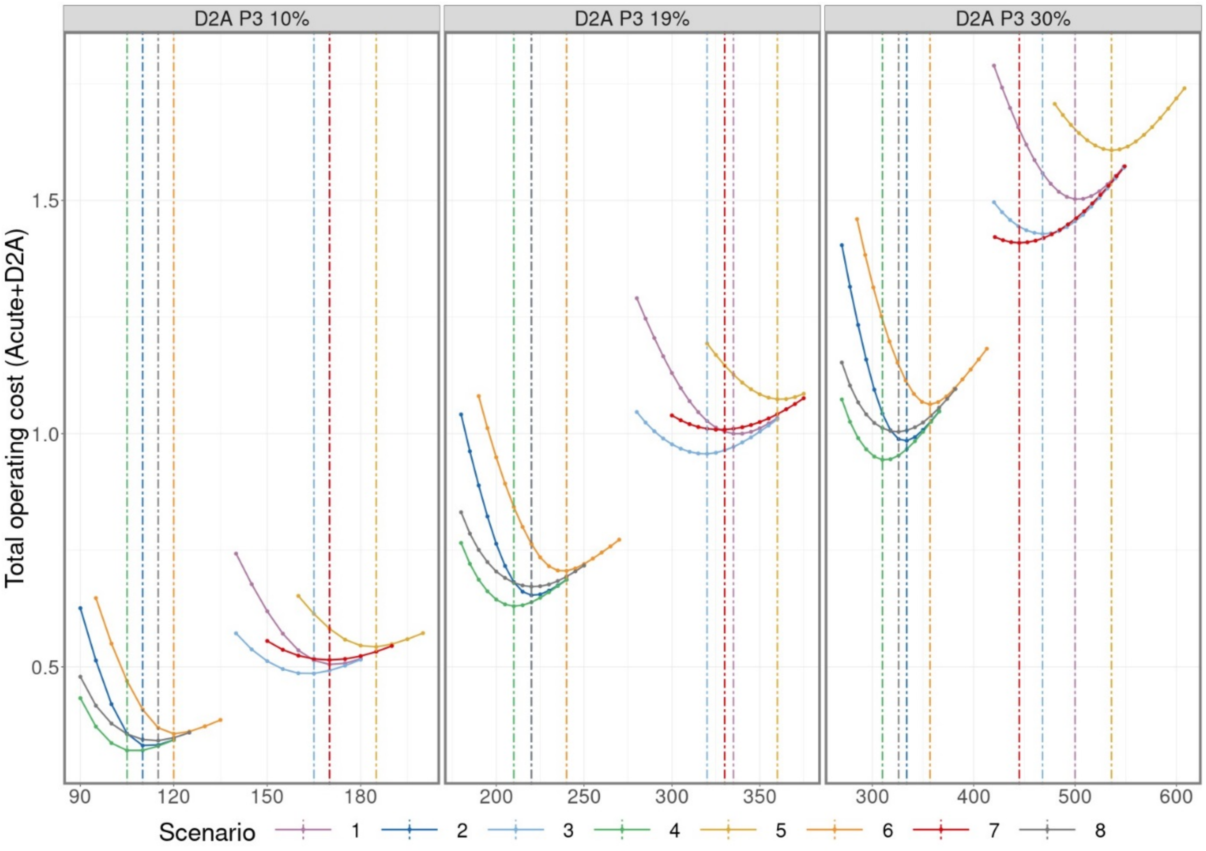


**Figure SM.B.3.** Weekly cost of acute delay and pathway care community service for capacity configurations for Pathway 3 with 10%, 19% (baseline) and 30% of acute admissions requiring complex community services at discharge.

### Supplementary Material C: Modelled results under assumption of no capacity constraint

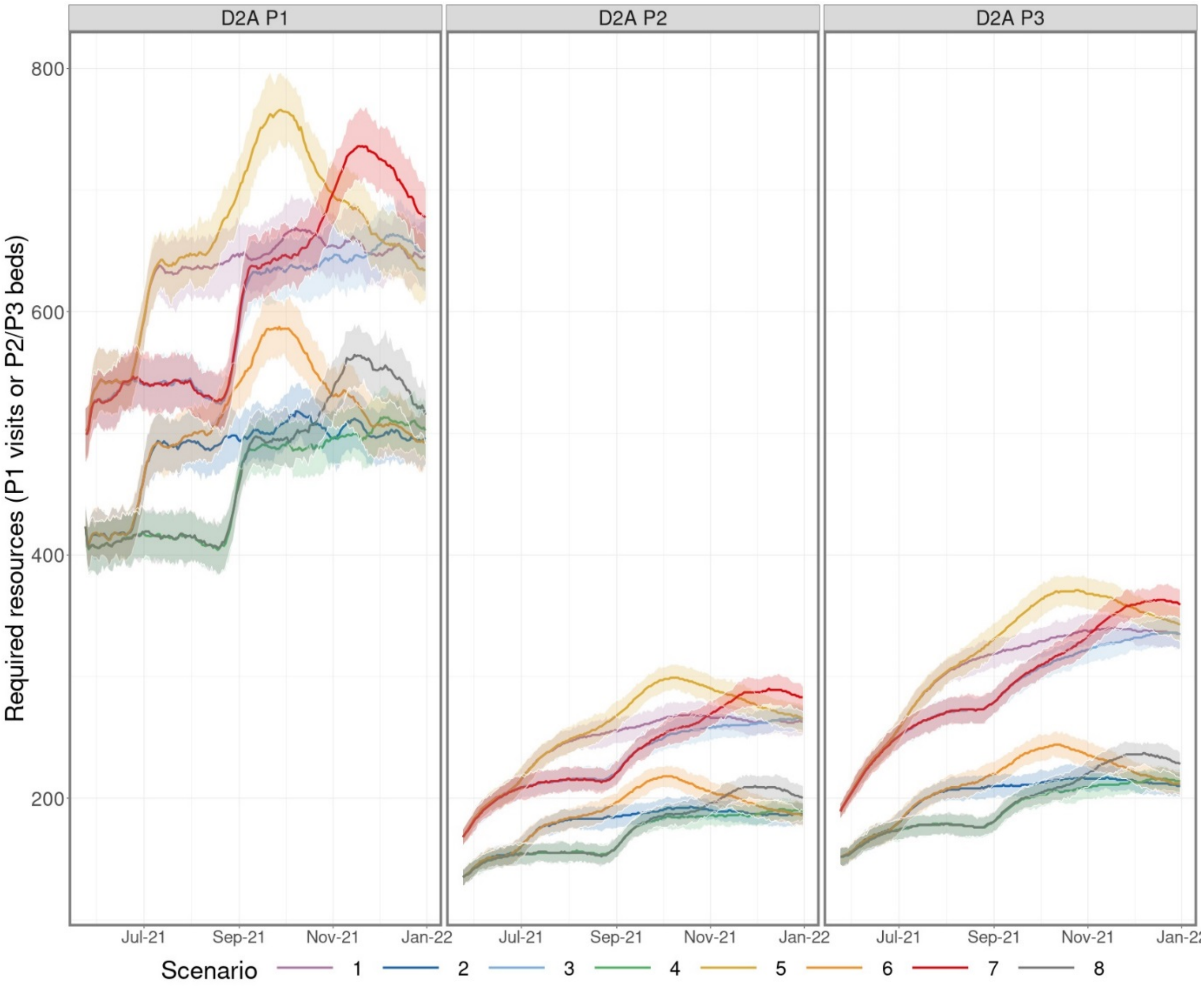


**Figure SM.C.1.** Projections of bed occupancy over time for considered scenarios under the assumption of no community services capacity constraint with interquartile range.

### Supplementary Material D: Research checklist (STRESS)

**Strengthening the Reporting of Empirical Simulation Studies (STRESS)**
**Discrete-event simulation guidelines STRESS-DES**

**Paper title:** Optimising the balance of acute and intermediate care capacity for the complex discharge pathway: computer modelling study during COVID-19 recovery in England

| Section/Subsection | Item | Recommendation | Submitted paper |
|---|---|---|---|
| **1. Objectives** | | | |
| Purpose of the model | 1.1 | Explain the background and objectives for the model. | To estimate the theoretical cost-optimal capacity requirement for 'step down' intermediate care services within a major healthcare system in England, at a time when considerable uncertainty remained regarding vaccination uptake and the easing of societal restrictions |
| Model Outputs | 1.2 | Define all quantitative performance measures that are reported, using equations where necessary. Specify how and when they are calculated during the model run along with how any measures of error such as confidence intervals are calculated. | Key simulation output measures of interest consist of: (i) Total cost of both intermediate care service provision (calculated from the modelled capacity levels) and the acute capacity required to support any delays to discharge (calculated from the mean number of acute beds blocked); (ii) Percentage occupancy per modelled capacity levels; (iii) Numbers of patients delayed per modelled capacity levels.<br><br>Each simulation was run until 31 December 2021, with 200 replications performed for each simulation in order to capture the realistic effect of variability (with respect to arrivals and lengths of stay). Results for each simulation were calculated from the outputs of these replications.<br><br>See also Methods section in paper and Supplementary Material A. |
| Experimentation Aims | 1.3 | If the model has been used for experimentation, state the objectives that it was used to investigate.<br><br>a.) Scenario based analysis – Provide a name and description for each scenario, providing a rationale for the choice of scenarios and ensure that item 2.3 (below) is completed. | Scenario based analysis. Full details of scenarios included in Table 1 in the main paper with explanations as to why each is investigated provided in the referring section (subsection Scenario Analysis within Methods chapter). |

| | | | |
|---|---|---|---|
| | | b.) Design of experiments – Provide details of the overall design of the experiments with reference to performance measures and their parameters (provide further details in *data* below).<br><br>c.) Simulation Optimisation – (if appropriate) Provide full details of what is to be optimised, the parameters that were included and the algorithm(s) that was be used. Where possible provide a citation of the algorithm(s). | |
| **2. Logic** | | | |
| Base model overview diagram | 2.1 | Describe the base model using appropriate diagrams and description. This could include one or more process flow, activity cycle or equivalent diagrams sufficient to describe the model to readers. Avoid complicated diagrams in the main text. The goal is to describe the breadth and depth of the model with respect to the system being studied. | 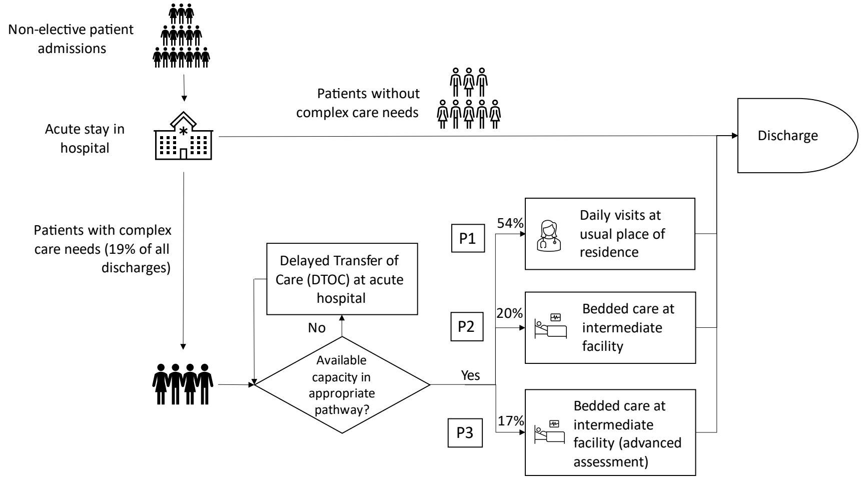  |
| Base model logic | 2.2 | Give details of the base model logic. Give additional model logic details sufficient to communicate to the reader how the model works. | See section Computer simulation modelling in chapter Methods and Supplementary Material A. |
| Scenario logic | 2.3 | Give details of the logical difference between the base case model and scenarios (if any). This could be | The difference between the base case model and the scenarios is in the values of the input parameters (clearly described in Table 1 in the main paper). |

| | | | | |
|---|---|---|---|---|
| | | incorporated as text or where differences are substantial could be incorporated in the same manner as 2.2. | | |
| Algorithms | 2.4 | Provide further detail on any algorithms in the model that (for example) mimic complex or manual processes in the real world (i.e. scheduling of arrivals/appointments/operations/maintenance, operation of a conveyor system, machine breakdowns, etc.). Sufficient detail should be included (or referred to in other published work) for the algorithms to be reproducible. Pseudo-code may be used to describe an algorithm. | | Implementation of this model is through the iterative three-phased method of discrete event simulation (Pidd, 1998).<br><br>Full details are provided in section Computer Simulation of the paper and Supplementary Material A. |
| Components | 2.5 | 2.5.1 Entities | Give details of all entities within the simulation including a description of their role in the model and a description of all their attributes. | Individual patients, each patient has an arrival time and a planned duration of care (either in bedded or visits based care) time as sampled from the appropriate length of stay distribution.<br><br>Full details are provided in section Computer Simulation of the paper and Supplementary Material A. |
| | | 2.5.2 Activities | Describe the activities that entities engage in within the model. Provide details of entity routing into and out of the activity. | Arrival (ready to be discharged from acute care hospital), start of service (visits or bedded depending on the pathway model) and end of service.<br><br>See Supplementary Material A for more details. |
| | | 2.5.3 Resources | List all the resources included within the model and which activities make use of them. | There are two types of resources depending on the pathway model: ‘slots’ for the visit-based care pathway model and ‘beds’ for the bedded care pathway model.<br><br>See Supplementary Material A for more details. |
| | | 2.5.4 Queues | Give details of the assumed queuing discipline used in the model (e.g. First in First Out, Last in First Out, prioritisation, etc.). Where one or more queues have a different discipline from the rest, provide a list of queues, indicating the queuing discipline used for each. If reneging, balking or jockeying occur, etc., provide | A simulated patient waiting to be scheduled in one of the three pathway models, is assumed to be occupying a bed in the acute care hospital.<br><br>Queue discipline is FIFO apart from the visit-based model (P1), where patients with a long duration of service, and/or a high initial/end visit requirement subsequently may prevent patients with lower service requirements from entering the P1 system. For this reason, if there are no available resources to start service for a new patient immediately, an arriving |

| | | | |
|---|---|---|---|
| | | details of the rules. Detail any delays or capacity constraints on the queues. | patient is scheduled to start on the following day. A patient whose visit sequence can be integrated into the available P1 capacity will be scheduled immediately.<br><br>See Supplementary Material A for more details. |
| | 2.5.5 Entry/Exit Points | Give details of the model boundaries i.e. all arrival and exit points of entities. Detail the arrival mechanism (e.g. 'thinning' to mimic a non-homogenous Poisson process or balking) | Entry: patient ready to be discharged from acute care hospital requiring placement in one of the three pathways (P1, P2, P3)<br>Exit point: discharged from one of P1, P2 or P3 |
| 3. **Data** | | | |
| Data sources | 3.1 | List and detail all data sources. Sources may include:<br><br>• Interviews with stakeholders,<br>• Samples of routinely collected data,<br>• Prospectively collected samples for the purpose of the simulation study,<br>• Public domain data published in either academic or organisational literature. Provide, where possible, the link and DOI to the data or reference to published literature.<br><br>All data source descriptions should include details of the sample size, sample date ranges and use within the study. | All secondary datasets used in the simulation models were supplied by the Clinical Commissioning Group (BNSSG CCG) and intermediate care providers (Sirona). Lengths of Stay distributions for each pathway were fitted to data 17/04/2020 – 10/02/2021 inclusive. At the start of each simulation, the initial occupancy and waiting list size was set equal to their actual values as of 14 May 2021, as provided by the CCG. Proportions of complex discharges and proportions of patients entering each pathway were empirically derived in collaboration with the CCG and detailed in Table SM.A.1. Cost sources are detailed in Table SM.A.2. |
| Pre-processing | 3.2 | Provide details of any data manipulation that has taken place before its use in the simulation, e.g. interpolation to account for missing data or the removal of outliers. | Demand projections are estimated from two different models, COVID-19 related demand from an SEIR model (Powell & Wood, 2021)., and the remaining demand from a regression model (Powell & Wood, 2021).<br><br>See sub-section Demand projection in the Methods section of the main paper. |
| Input parameters | 3.3 | List all input variables in the model. Provide a description of their use and include parameter values. For stochastic inputs provide details of any continuous, discrete or empirical distributions used | See Tables SM.A.1 and 2 in the Supplementary Material A. |

| | | | |
|---|---|---|---|
| | | along with all associated parameters. Give details of all time dependent parameters and correlation.<br><br>Clearly state:<br><br>• Base case data<br>• Data use in experimentation, where different from the base case.<br>• Where optimisation or design of experiments has been used, state the range of values that parameters can take.<br><br>Where theoretical distributions are used, state how these were selected and prioritised above other candidate distributions. | |
| Assumptions | 3.4 | Where data or knowledge of the real system is unavailable what assumptions are included in the model? This might include parameter values, distributions or routing logic within the model. | See assumptions in Methods section and limitations in the Discussion. |
| 4. **Experimentation** | | | |
| Initialisation | 4.1 | Report if the system modelled is terminating or non-terminating. State if a warm-up period has been used, its length and the analysis method used to select it. For terminating systems state the stopping condition.<br><br>State what if any initial model conditions have been included, e.g., pre-loaded queues and activities. Report whether initialisation of these variables is deterministic or stochastic. | Non-terminating system. No warm-up period was used. Instead, at the start of each simulation, the initial occupancy and waiting list size was set equal to their actual values as of 14 May 2021.<br><br>Full details are provided in section Computer Simulation of the paper and Supplementary Material A. |
| Run length | 4.2 | Detail the run length of the simulation model and time units. | Time unit is day. Simulation runs until 31 Dec 2021. |

| | | | |
|---|---|---|---|
| Estimation approach | 4.3 | State the method used to account for the stochasticity: For example, two common methods are multiple replications or batch means. Where multiple replications have been used, state the number of replications and for batch means, indicate the batch length and whether the batch means procedure is standard, spaced or overlapping. For both procedures provide a justification for the methods used and the number of replications/size of batches. | 200 multiple replications were used for each scenario. |
| 5. **Implementation** | | | |
| Software or programming language | 5.1 | State the operating system and version and build number.<br><br>State the name, version and build number of commercial or open source DES software that the model is implemented in.<br><br>State the name and version of general-purpose programming languages used (e.g. Python 3.5).<br><br>Where frameworks and libraries have been used provide all details including version numbers. | The model was coded from scratch in R and has been released as an open-source tool (hosted on https://github.com/nhs-bnssg-analytics/ipacs-v1-model and promoted via social media).<br><br>Model data available at https://github.com/nhs-bnssg-analytics/c19-recovery-community-services-modelling |
| Random sampling | 5.2 | State the algorithm used to generate random samples in the software/programming language used e.g. Mersenne Twister.<br><br>If common random numbers are used, state how seeds (or random number streams) are distributed among sampling processes. | Uses the inbuilt random number generator in R. Each replication uses a different seed call to this function. This provides the necessary stochastic variation within each replication, yet also allows reproducible model scenarios to be created and assessed (useful when evaluating specific changes in the model parameters). |
| Model execution | 5.3 | State the event processing mechanism used e.g. three phase, event, activity, process interaction. | In this study, 'discrete time' simulation was used to dynamically model the flow of individual patients from acute discharge readiness (i.e. to become an 'arrival' at the start of a D2A waiting list) through to completion of intermediate care. |

| | | | |
|---|---|---|---|
| | | *Note that in some commercial software the event processing mechanism may not be published. In these cases authors should adhere to item 5.1 software recommendations.*<br><br>State all priority rules included if entities/activities compete for resources.<br><br>If the model is parallel, distributed and/or use grid or cloud computing, etc., state and preferably reference the technology used. For parallel and distributed simulations the time management algorithms used. If the HLA is used then state the version of the standard, which run-time infrastructure (and version), and any supporting documents (FOMs, etc.) | Essentially this involves simulating the arrival of simulated individual patients (see Method; Demand projection), commencement of intermediate care in the relevant D2A pathway (for which the patient may have to wait, depending on available capacity), and their departure from the service (determined by their length of stay).<br><br>A separate model was constructed for each of the three D2A pathways and the eight scenarios considered (Table 1). At the start of each simulation, the initial occupancy and waiting list size was set equal to their actual values as of 14 May 2021. Within each simulation, each future day (the 'discrete time' interval used in this study) was simulated consecutively, with instances of the above-mentioned three events performed in line with the simulation schedule (the arrivals onto the D2A pathway which were due to occur that day; how many patients were due to complete their D2A pathway that day; and the commencement of intermediate care provided there were patients waiting for care and available capacity).<br><br>More information in Supplementary Material A. |
| System Specification | 5.4 | State the model run time and specification of hardware used. This is particularly important for large scale models that require substantial computing power. For parallel, distributed and/or use grid or cloud computing, etc. state the details of all systems used in the implementation (processors, network, etc.) | Processing time is insubstantial, typically taking less than five minutes for each scenario evaluated on a desktop computer. |
| **6. Code Access** | | | |
| Computer Model Sharing Statement | 6.1 | Describe how someone could obtain the model described in the paper, the simulation software and any other associated software (or hardware) needed to reproduce the results. Provide, where possible, the link and DOIs to these. | The tool is open source and available for free: https://github.com/nhs-bnssg-analytics/ipacs-v1-model |
